# Penetrance of breast cancer genes from the eMERGE III Network

**DOI:** 10.1101/2021.04.24.21255936

**Authors:** Xiao Fan, Julia Wynn, Ning Shang, Cong Liu, Alexander Fedotov, Miranda L.G. Hallquist, Adam H. Buchanan, Marc S. Williams, Maureen E. Smith, Christin Hoell, Laura J. Rasmussen-Torvik, Josh F. Peterson, Georgia L. Wiesner, Andrea M. Murad, Gail P. Jarvik, Adam S. Gordon, Elisabeth A. Rosenthal, Ian B. Stanaway, David R. Crosslin, Eric B. Larson, Kathleen A. Leppig, Nora B. Henrikson, Janet L. Williams, Rongling Li, Scott Hebbring, Chunhua Weng, Yufeng Shen, Katherine D. Crew, Wendy K. Chung

## Abstract

We studied the penetrance and clinical outcomes of seven breast cancer susceptibility genes (*BRCA1, BRCA2, TP53, CHEK2, ATM, PALB2* and *PTEN*) in almost 25,000 participants unselected for personal or family history of breast cancer. We identified 420 participants with pathogenic or likely pathogenic variants, and 147 were women who did not previously know their genetic results. Out of these 147 women, 32 women were diagnosed with breast cancer at an average age of 52.8 years. Estimated penetrance by age 60 years ranged from 18-44%, depending on the gene. Within the first twelve months after genetic results disclosure, 42% of women had taken actions related to their genetic results and two new breast cancer cases were identified. Our study provides population-based penetrance estimates for the understudied genes, *CHEK2, ATM*, and *PALB2*, and highlights the importance of using unselected populations for penetrance studies. It also demonstrates the potential clinical impact of genetic testing to improve healthcare through early diagnosis and preventative screening.

## Introduction

Breast cancer is the most common cancer in women after skin cancer, annually affecting 2.1 million women worldwide with an estimated 627,000 annual deaths [1]. Approximately 5-10% of breast cancer cases are associated with a highly penetrant monogenic variant and are genetically heterogenous [2-6]. With the exception of a small number of founder pathogenic variants, the majority of pathogenic variants in hereditary cancer genes are very rare or unique to the individual tested [7]. Multiple studies of cancer penetrance for hereditary breast cancer genes have been conducted since the identification of *BRCA1* and *BRCA2* in 1994 and 1995, respectively [8, 9]. Early penetrance studies were often enriched for participants with strong family histories of breast cancer or early age of onset of breast cancer [10, 11]. Recent studies have highlighted that penetrance estimates are dependent on the method of ascertainment, and disease risks of familial cases are higher than risk estimates derived from the general population [12-14]. In the Breast Cancer Linkage Consortium (BCLC) families, penetrance of *BRCA1* and *BRCA2* by age 70 years was estimated as 85% [15] and 84% [10], respectively. However, a population-based study estimated penetrance of 52% for *BRCA1* and 32% for *BRCA2* by age 70 years [16]. Unbiased estimates of penetrance are challenging, but are critically important to make informed choices about strategies for risk reduction through increased surveillance and risk reducing interventions including prophylactic surgery.

A few studies have estimated the penetrance of *BRCA1*/2 pathogenic variants in the general population [16-18], which require a large number of individuals to be sequenced in order to identify a meaningful number of individuals who carry pathogenic variants. Population-based penetrance estimates are not available for the other hereditary breast cancer genes with lower frequency of pathogenic variants (*e*.*g*., *TP53* and *PTEN*), more recently identified genes (*e*.*g*., *PALB2*), and those that likely have more moderate penetrance (*e*.*g*., *ATM* and *CHEK2)*. Increasingly, there is consideration of population-based genomic health screening for adults for conditions for which surveillance is effective, including breast cancer [19]. *BRCA*-associated hereditary breast and ovarian cancer is classified by the Centers for Disease Control and Prevention as tier 1 application for interventions that reduce morbidity and mortality in individuals with certain personal and family history indications [20]. Unbiased estimates of risk are necessary to accurately estimate risk over the life course if population-based screening is deployed. Our study objective is to provide less biased estimates of breast cancer penetrance in women for the commonly assessed breast cancer genes [21] in clinical genetic testing (*e*.*g*., *BRCA1, BRCA2, PALB2, PTEN, TP53, ATM* and *CHEK2*).

## Methods

### Study cohort and sequencing panel

In phase III of the Electronic Medical Records and Genomics (eMERGE) Network, 24,947 participants were enrolled at 11 clinical sites and had sequencing for a panel of 109 genes [22]. The focus of this analysis is seven breast cancer susceptibility genes included on the eMERGE III panel (*BRCA1, BRCA2, PALB2, PTEN, TP53, ATM*, and *CHEK2*). Variants identified through sequencing were classified according to the American College of Medical Genetics and Genomics and Association for Molecular Pathology (ACMG-AMP) guidelines [23, 24]. Pathogenic or likely pathogenic (P/LP) variants were confirmed by Sanger sequencing [22]. We focused on female participants in this study as the risk for breast cancer in men with P/LP variants is significantly lower than that for women. Putative somatic variants (variant allele fraction in blood samples <0.3) were excluded from downstream analysis. Participants who previously knew their genetic results may be more likely to have a high risk for breast cancer, have a known genetic risk in their family, and to have undergone risk reduction or enhanced surveillance actions after they received their genetic results. Alternatively, we assumed that the participants who were unaware of their genetic risk are more likely to have a breast cancer prevalence similar to the general population risk. There only women with P/LP variants who were unaware of the genetic risk were included in the pentrance and clinical impact analysis. For clinical impact analysis, the sample was further restricted to those participants who consented to return of results (RoR) [25] and did not have breast cancer before RoR. We assessed clinical impact at 12 months post-RoR. All 11 clinical sites consented participants under institutional review board-approved protocols [26].

### Identification of breast cancer diagnosis, and post-RoR risk management

One of the major goals of eMERGE III was to study the return of actionable genetic variants to participants and assess impact on clinical care. The electronic health records (EHR) of participants with breast cancer susceptibility P/LP variants were manually queried at twelve months post-RoR for any history of incident breast cancer and the date recorded in the EHR to ascertain if the event occurred prior to or after results disclosure. We measured the post-RoR performance of risk management including breast cancer surveillance and prevention procedures such as breast magnetic resonance imaging (MRI), breast ultrasound, mammograms, breast cancer risk-reducing medication, and prophylactic mastectomy and oophorectomy. We also recorded breast biopsies as a diagnostic test. Records were queried for prior patient knowledge of the identified breast cancer susceptibility P/LP variant. EHR extraction was completed at each clinical site and entered into a central REDCap database [27, 28].

### Penetrance estimation

We used Kaplan-Meier method [29] to estimate the age-specific cumulative risk of breast cancer in the women with P/LP variants in each breast cancer gene. The participants were censored at their current age or age of prophylactic risk reduction surgery (mastectomy or oophorectomy). We re-estimated penetrance whenever an event occurred in the curve of cumulative risk of breast cancer. We also estimated breast cancer penetrance by decade from 30 to 70 years. Confidence intervals were calculated using Greenwood’s formula [30]. The analysis was done in R version 3.6.3.

## Results

### Clinical characteristics

Of the 24,947 eMERGE III participants who were sequenced, 13,458 were women (Supplementary Table S1). We identified 420 individuals with at least one P/LP variant in one of the seven breast cancer susceptibility genes. A flowchart of inclusion criteria for participants is shown in Figure 1. Although we focused on penetrance of breast cancer in women, we identified 169 men with P/LP variants in the seven breast cancer genes, and one man (*BRCA2*: c.7758G>A) out of 55 men with *BRCA2* P/LP variants developed breast cancer. We excluded nine women with putative somatic variants and 99 women who already knew their genetic results, and retained 147 women for the penetrance analysis. The clinical characteristics of this female cohort are shown in Table 1 and Supplementary Figure S1. The majority (73%) were of European and non-Latinx ancestry, followed by 16% African American, 9% Latinx and 3% East Asian by self-reported ancestry. The average age was 55.1 years (standard deviation of 18.2 years). One woman had two P/LP variants including one frameshift variant in *BRCA2* and one missense variant in *CHEK2*. By the date of last chart review, 32 (21.8%) women had developed breast cancer and two of them were diagnosed post-RoR. The average age of breast cancer diagnosis was 52.8 years (95% CI: 30.8-74.8), and four (2.7%) women had a prophylactic mastectomy or oophorectomy after eMERGE RoR. While not statistically significant (*P*-value=0.11), by age 50 years, a higher proportion of African American (7 out of 13, 53.8%) and Latinx (5 out of 11, 45.5%) individuals developed breast cancer compared with European-ancestry individuals (20 out of 72, 27.8%), using the binomial test.

**Table 1.**
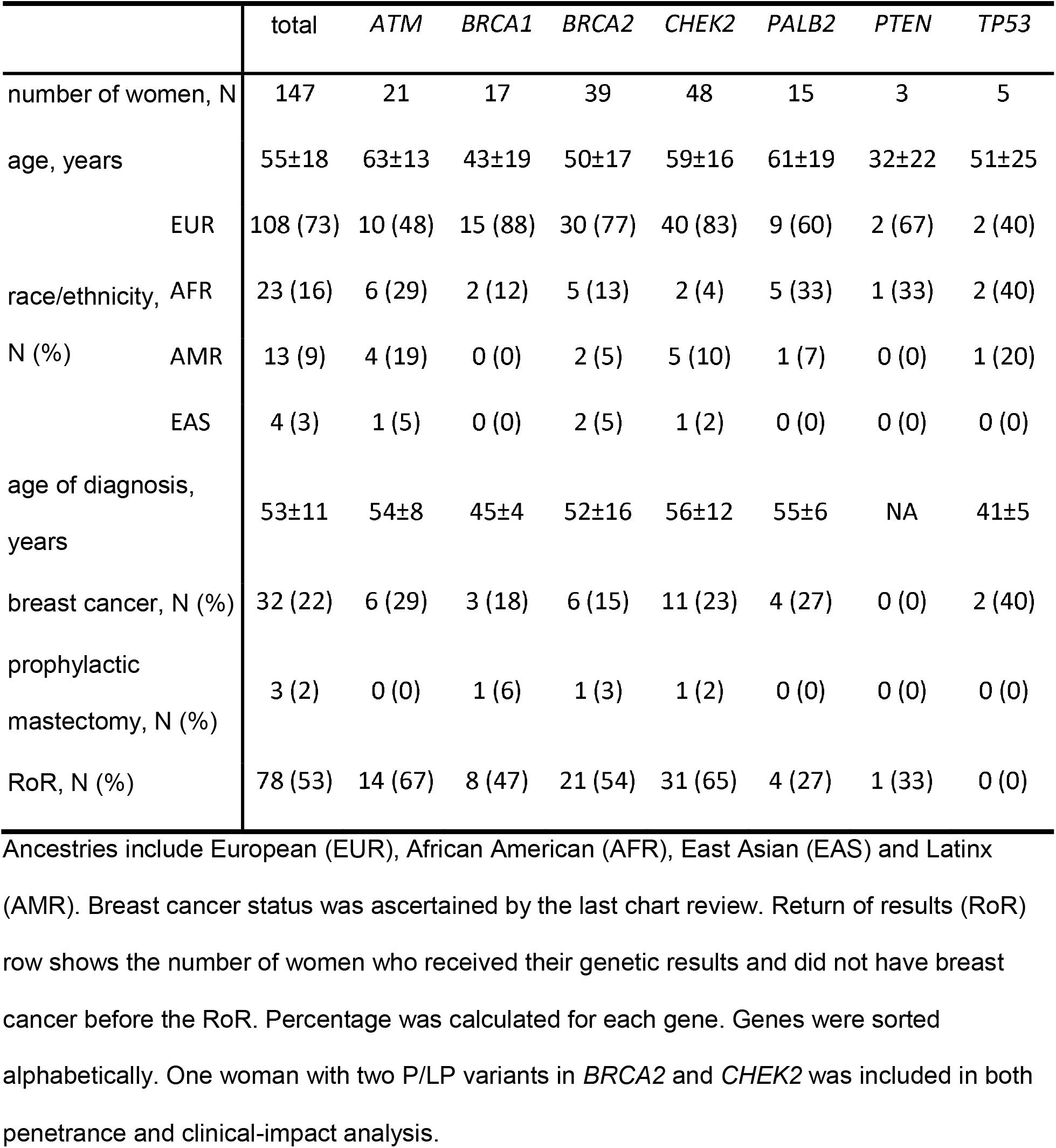
**Clinical characteristics of 147 women with germline pathogenic/likely pathogenic variants in the seven breast cancer genes.**

**Figure 1.**
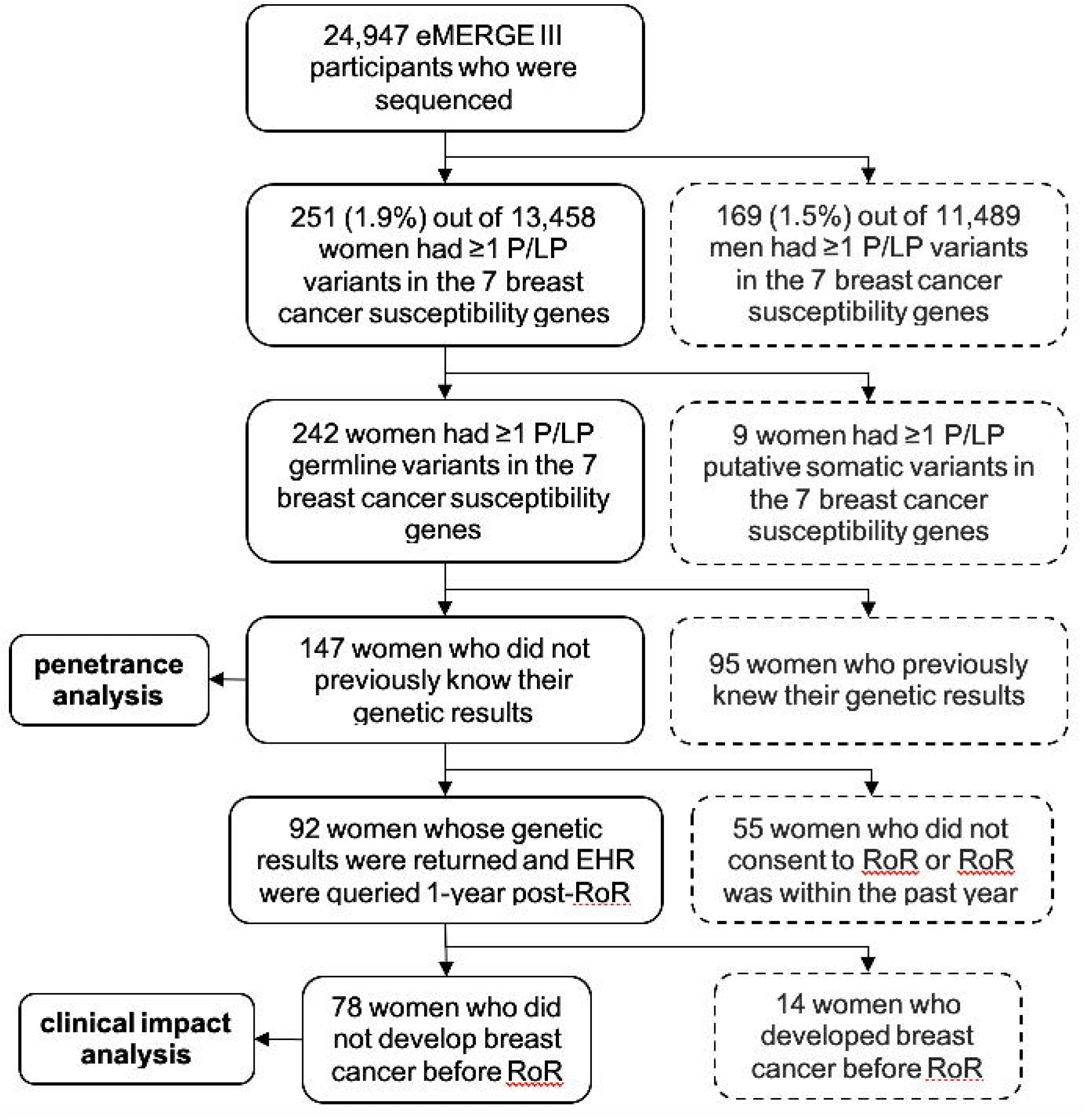
Study cohorts for penetrance analysis and clinical impact analysis. Participants in the dashed box were excluded.

**Figure 2.**
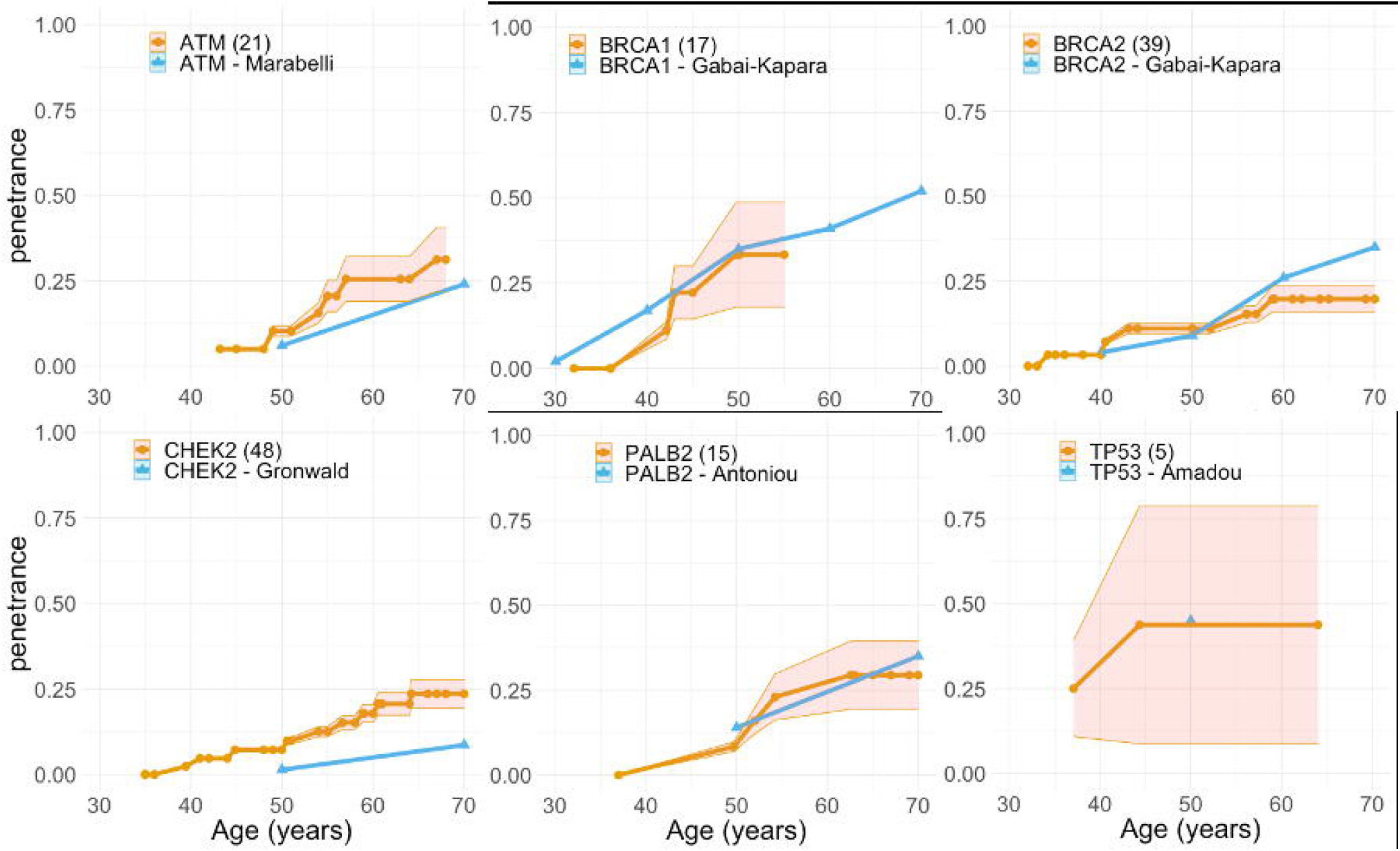
**Breast cancer penetrance (orange) of six breast cancer genes compared with penetrance (blue) in the literature from Supplementary Table S2. The sample size of the eMERGE III penetrance cohort was annotated in the parentheses after the gene in the legend. Orange bands indicate 95% confidence intervals. Genes were sorted alphabetically.**

### Variant characteristics

The 147 women had 100 unique P/LP variants in the seven breast cancer susceptibility genes. Fourteen variants were recurrent in this cohort (Supplementary Table S2), the most frequent of which were *CHEK2* c.470T>C (n=26), an Ashkenazi

Jewish founder variant, and *CHEK2*: c.1100delC (n=7), a European founder variant. The most commonly reported variant type in this cohort was putative loss of function (LoF) including frameshift, stop-gain, or splice for *BRCA2, BRCA1, ATM*, and *PALB2. CHEK2* had both P/LP missense and putative LoF variants beside two deletions.

Compared with 90% of 85 putative LoF variants that were annotated in ClinVar with at least two stars in review status (assertion criteria provided, multiple submitters and no conflicts), fewer (62% of 13) missense variants were well documented in ClinVar. The numbers of women with P/LP variants and the numbers of unique P/LP variants for each gene are shown in the Supplementary Table S3. For *ATM, BRCA1, BRCA2*, and *PALB2*, over 80% of their P/LP variants were annotated in ClinVar with two or three stars in review status. However, out of 13 P/LP *CHEK2* variants in 48 women, only seven (54%) of the variants in seven (15%) women have been well annotated in ClinVar.

### Prevalence and penetrance

We identified 420 (1.7%) of 24,947 eMERGE participants with at least one P/LP variant among the seven breast cancer susceptibility genes, with *BRCA1/2* P/LP variants being most commonly identified (225 (0.9%) participants). Removing individuals who previously knew their genetic results, 137 (0.5%) unselected individuals had *BRCA1/2* P/LP variants. This prevalence of 0.5% is consistent with prevalence of *BRCA1/2* P/LP variants reported in a previous study [31]. The frequency of *BRCA1/2* P/LP variants differ across race and ethnicity. In the cohort of 147 women, 37.1.9% of them had P/LP variants in *BRCA1* or *BRCA2*. When stratified, the frequency is 41.7% in the 108 European-ancestry individuals, 30.4% in the 23 African Americans, 15.4% in the 13 Latinx, and 50% in the four Asians. *CHEK2, ATM*, and *PALB2* P/LP variants accounted for 32.7%, 14.3% and 10.2%, respectively, of those 147 women.

Overall, *BRCA1* and *TP53* had high breast cancer penetrance and included early-onset breast cancer diagnosed before age 50. *BRCA2, ATM, PALB2*, and *CHEK2* were more commonly associated with later onset breast cancer, for which the penetrance estimates ranged from 19% to 31% by age 60 years (Table 2). We compared the penetrance estimated using the eMERGE III data and penetrance reported from previous studies in Figure 1. Supplementary Table 4 summarizes published penetrance studies for seven breast cancer genes. Excluding the individuals who knew their genetic results for breast cancer genes, the penetrance of our study cohort is close to that of the general population unselected for personal history of breast cancer. For comparison, we calculated the penetrance estimates using all 242 women including those who previously knew their genetic results in Supplementary Figure S2. As expected, the penetrance estimated including women with prior knowledge of a hereditary breast cancer susceptibility, for whom genetic testing were usually performed after a diagnosis of cancer, is higher than the penetrance estimated using unselected women.

**Table 2.** Penetrance by decades of six breast cancer genes.

|  | <30 years | <40 years | <50 years | <60 years | <70 years |
| --- | --- | --- | --- | --- | --- |
| *ATM* | 0.0 [0.0, 0.0] | 0.0 [0.0, 0.0] | 10.3 [8.7, 11.8] | 25.5 [18.9, 32.1] | 31.2 [21.8, 40.7] |
| *BRCA1* | 0.0 [0.0, 0.0] | 0.0 [0.0, 0.0] | 33.3 [17.9, 48.7] | 33.3 [17.9, 48.7] | 33.3 [17.9, 48.7] |
| *BRCA2* | 0.0 [0.0, 0.0] | 3.3 [3.1, 3.6] | 11.1 [9.6, 12.5] | 19.8 [15.9, 23.6] | 19.8 [15.9, 23.6] |
| *CHEK2* | 0.0 [0.0, 0.0] | 2.4 [2.3, 2.5] | 7.2 [6.6, 7.8] | 17.8 [15.2, 20.4] | 23.6 [19.4, 27.8] |
| *PALB2* | 0.0 [0.0, 0.0] | 0.0 [0.0, 0.0] | 8.3 [6.9, 9.8] | 23.0 [16.2, 29.8] | 29.4 [19.4, 39.4] |
| *TP53* | 0.0 [0.0, 0.0] | 25.0 [10.9, 39.1] | 43.8 [8.7, 78.8] | 43.8 [8.7, 78.8] | 43.8 [8.7, 78.8] |
| general population* |  |  | 2.0 | 4.4 | 7.7 |

When the number of uncensored women who had P/LP variants is below five (sample size is too small), the penetrance estimate is not accurate. We annotated these penetrance estimates in orange. \*risk of breast cancer in populations was obtained from Cancer Statistics Center.

### ATM

Twenty-one women had *ATM* P/LP variants, and 18 of them carried putative LoF variants. Six women developed breast cancer at an average age of 54.2 years. Three *ATM* variants were recurrent. Two women with the same missense variant *ATM*: c.7271T>G and one woman with a different missense variant *ATM*: c.6095G>A all developed breast cancer at a young average age of 48.7 years. The estimated penetrance overall was 5% by age 50 which is similar to the 6% reported by Marabelli *et al*. [32].

### BRCA1

We identified 17 women with *BRCA1* P/LP variants. The penetrance estimate of 33% by age 50 in this eMERGE cohort is comparable with that previously reported of 35% by Gabai-Kapara *et al*. [16]. However, our *BRCA1* cohort is too young to have accurate penetrance estimates beyond 60 years of age with only one woman not censored by age 60. Women with *BRCA1* P/LP variants had early onset of breast cancer at an average age of 45.0 years.

### BRCA2

There were 39 women with *BRCA2* P/LP variants. The number is twice of that for *BRCA1*, a ratio consistent with previous findings [31, 33]. All reported P/LP *BRCA2* variants in the eMERGE cohort were putative LoF. Six of the 39 women had a history of breast cancer at an average age of 51.7 years. The penetrance estimate to age 60 is 20% which is slightly lower than 26% reported by Gabai-Kapara *et al*. [16].

### CHEK2

*CHEK2* P/LP variants were identified in 48 women in the eMERGE cohort. Twenty-six of the women carried the Jewish founder missense variant c.470T>C. This variant is present in 0.46-1.2% of the Ashkenazi Jewish population [34, 35] and is a moderate risk variant associated with a 1.5 fold relative risk for breast cancer. The most common European founder variant c.1100delC was found in seven women and was previously reported to have a penetrance of 37% by age 70. We do not have sufficient sample size to estimate penetrance of this single variant, but of four women in our cohort, one of them developed breast cancer by age 50. Though the penetrance of *CHEK2* is moderate, our estimate is higher than that previously reported. By age 75 years, the penetrance of 27.7% (95% CI: 21.7%, 32.7%) estimated in the eMERGE III cohort was significantly higher than 10.4% reported in the Gronwald *et al*. [36].

### PALB2

Out of 15 women with *PALB2* P/LP variants, four had breast cancer at an average age of 50.6 years. All P/LP *PALB2* variants were putative LoF. Our estimated penetrance of 8% is slightly lower than that reported by Antoniou *et al*. [37] of 14% by age 50 years.

### PTEN

We identified three women with P/LP variants in *PTEN*. None of the women had breast cancer with the current ages of 15, 24 and 57 years. The penetrance of *PTEN* was not further analyzed due to the limited sample size.

### TP53

Of the five women with *TP53* P/LP variants, three of them had missense variants. Two women with missense variants developed breast cancer at the ages of 37.1 and 44.4 years. Though the sample size is small, our penetrance estimate of 44% is consistent with the reported penetrance of 45% by age 50 years [38].

### Clinical impact

Among the 242 women with P/LP variants in the seven breast cancer susceptibility genes, not all of them consented to receive their genetic results. We determined that 183 genetic results were returned to female participants and 91 (50.0%) women previously knew their genetic results, most of which (90.5%) were *BRCA1* or *BRCA2* variants. We limited the analysis of clinical impact after RoR to the 78 women not previously aware of the genetic results and without a breast cancer diagnosis before RoR. Based on the chart review at twelve-month post-RoR, 26 women had mammograms, 11 had breast MRI, five had breast ultrasound. Three women had breast biopsies which led to the diagnosis of a new breast cancer in two women. From the perspective of risk reduction, three women had prophylactic bilateral mastectomies and one also had a prophylactic oophorectomy, and another woman had only a prophylactic oophorectomy. One woman started tamoxifen to reduce breast cancer risk. Overall, 33 (42.3%) women took one or more clinical actions, not including the breast biopsies, and 9 (11.5%) had at least two breast cancer surveillance procedures after RoR.

## Discussion

In this study, we investigated penetrance of breast cancer in seven high or moderate-penetrance breast cancer susceptibility genes (*BRCA1, BRCA2, TP53, CHEK2, ATM, PALB2*, and *PTEN*) using the eMERGE III cohort. We identified 147 women with P/LP genetic variants in the seven genes, who represent a population unselected for personal or family history of breast cancer. Across the genes, *BRCA1* and *TP53* variants were associated with high penetrance, and variants in *BRCA2, ATM, PALB2*, and *CHEK2* had more moderate penetrance. Even the moderate-penetrance genes predisposed a significantly higher risk of breast cancer than average-risk women (Table 2) [39]. Our findings suggest that *CHEK2* P/LP variants could be as prevalent as *BRCA1*/2 in certain populations such as those of Ashkenazi Jewish or European ancestry with *CHEK2* founder variants. We estimated higher penetrance for *CHEK2* compared with estimates from a prior study [36]. Prospective data from larger numbers of participants will help to better estimate penetrance and could impact breast cancer surveillance for women with *CHEK2* P/LP variants.

We highlight the importance of estimating penetrance using unselected populations by comparing penetrance estimates before and after removing women who knew their genetic results prior to enrollment in eMERGE. Penetrance of *BRCA1/2* increased dramatically when women who knew their genetic result were included, suggesting use of selected populations inflates penetrance estimates.

Successful impact of population screening relies on uptake of surveillance for early detection (such as breast MRI), risk reducing surgeries and chemoprevention for primary prevention of breast cancer. The National Comprehensive Cancer Network (NCCN) provides guidelines for breast cancer risk management to women with P/LP variants conferring high risk for breast cancer [40, 41]. The seven genes we studied are all included in the NCCN suggested breast cancer gene panel for cancer risk management. For women who desire risk-reducing interventions, risk-reducing medications (such as selective estrogen receptor modulators and aromatase inhibitors) and surgeries (prophylactic mastectomies and oophorectomies) are considerations.

Routine genetic screening could lead to improvement in long-term clinical outcomes with tailored health surveillance. Among 242 women with P/LP variants in the seven genes, 147 (61%) of them were not previously aware of their genetic results. This research supports the feasibility of identification of women at increased risk of breast cancer on a population level.

Among 74 women who first learned their genetic results through eMERGE, 42% of women had taken actions related to their genetic results within 12 months, though the uptake of mammograms (33.3%), breast MRI (14.1%) and mastectomy (3.8%) was lower than that found in a study of women with *BRCA1/2* P/LP variants in which uptake of the same actions was 45.8%, 32.2% and 3.5%, respectively within the first year post RoR [31]. In contrast to eMERGE where there was variability across sites for the RoR process [25], this was a single site study with an established infrastructure to support participants after the return of results [42]. Besides, we only assessed clinical actions through the EHR at the eMERGE participating institutions, and it is possible clinical actions were taken by participants at other clinical sites and not captured in the EHR we queried. With this small sample size of 74 women, two new breast cancer cases were diagnosed within the first twelve months after RoR, demonstrating the potential impact of returning this information.

## Limitations

It is challenging to recruit a large, unbiased cohort from the general population. Although eMERGE III had ∼25,000 participants sequenced, the number of women with P/LP variants in breast cancer genes is small compared with studies selected for family history. Unbiased penetrance estimates using larger sample sizes are still needed.

Breast cancer is caused by a combination of genetic, hormonal, and environmental factors. We focused on some genetic factors in this study, and other factors such as family history, breast density, reproductive history and body mass index were not considered.

The genes we studied are commonly tested clinically when assessing breast cancer risk [21]. There are other genes that are less commonly associated with breast cancer that are included with comprehensive clinical breast cancer genetic assessments including *CDH1, STK11, NBN*, and *NF1* that we did not include in the eMERGE III gene panel due to the low prevalence and unclear penetrance. Future studies with much larger sample sizes are needed to estimate penetrance for genes with low population prevalence.

A limitation of our clinical impact study was that a large portion of women did not consent to receive their genetic results, were already aware of their results prior to participation in eMERGE III, or had developed breast cancer before RoR, limiting the sample size for this analysis. We only have access to data from the EHR at the eMERGE sites, therefore clinical actions performed outside of the eMERGE sites was not assessed. One-year of follow-up may be insufficient time to assess impact, but we were limited by the project period. We did not assess lifestyle modifications adopted by participants to reduce cancer risk such as maintaining a healthy body weight, moderate physical activity, and reducing alcohol consumption.

## Conclusions

We studied the penetrance of seven commonly tested breast cancer susceptibility genes in a cohort that was unselected for personal or family history of breast cancer. Our study highlights the importance of unbiased recruitment for estimates of cancer risk as we consider population-based testing and risk stratification. Population-based penetrance estimates based on larger sample sizes for *CHEK2, ATM*, and *PALB2* will be useful for genetic counseling for breast cancer risk and subsequent recommendations for cancer risk management. Genetic testing for breast cancer genes including the moderate-penetrance genes in healthy population has the potential to diagnose cancers at earlier more treatable stages with enhanced surveillance, and perform risk reducing surgeries such as prophylactic mastectomies and oophorectomies. We demonstrate that genetic screening for breast cancer genes can lead to early diagnosis and potentially prevent cancer.

## Data Availability

All data sets summarized here will be publicly available in the dbGaP repository under phs001616.v1.p1 and pre-dbGaP submission access can also be requested on the eMERGE website.

https://emerge-network.org

## Acknowledgements

The authors wish to acknowledge the participating patients and family who generously contributed to the eMERGE III data. We thank the participating sites and coordinating centers for the participant recruitment and follow-up data collection.

## Notes

Murad, AM is a paid consultant for Concert Genetics. The other authors have no conflicts of interest to disclose.

## Data Availability Statement

All data sets summarized here will be publicly available in the dbGaP repository under phs001616.v1.p1 and pre-dbGaP submission access can also be requested on the eMERGE website https://emerge-network.org.

## References

1. World Health Organization, Breast Cancer. 2019; Available from: https://www.who.int/cancer/prevention/diagnosis-screening/breast-cancer/en/.

2. Claus, E.B., N. Risch, and W.D. Thompson, Genetic analysis of breast cancer in the cancer and steroid hormone study. Am J Hum Genet, 1991. 48(2): p. 232–42.

3. Hall, J.M., et al., Linkage of early-onset familial breast cancer to chromosome 17q21. Science, 1990. 250(4988): p. 1684–9.

4. Newman, B., et al., Inheritance of human breast cancer: evidence for autosomal dominant transmission in high-risk families. Proc Natl Acad Sci U S A, 1988. 85(9): p. 3044–8.

5. Miki, Y., et al., A strong candidate for the breast and ovarian cancer susceptibility gene BRCA1. Science, 1994. 266(5182): p. 66–71.

6. Prevalence and penetrance of BRCA1 and BRCA2 mutations in a population-based series of breast cancer cases. Anglian Breast Cancer Study Group. Br J Cancer, 2000. 83(10): p. 1301–8.

7. Neben, C.L., et al., Multi-Gene Panel Testing of 23,179 Individuals for Hereditary Cancer Risk Identifies Pathogenic Variant Carriers Missed by Current Genetic Testing Guidelines. J Mol Diagn, 2019. 21(4): p. 646–657.

8. Narod, S.A. and W.D. Foulkes, BRCA1 and BRCA2: 1994 and beyond. Nat Rev Cancer, 2004. 4(9): p. 665–76.

9. Wooster, R., et al., Identification of the breast cancer susceptibility gene BRCA2. Nature, 1995. 378(6559): p. 789–92.

10. Ford, D., et al., Genetic heterogeneity and penetrance analysis of the BRCA1 and BRCA2 genes in breast cancer families. The Breast Cancer Linkage Consortium. Am J Hum Genet, 1998. 62(3): p. 676–89.

11. Meijers-Heijboer, H., et al., Low-penetrance susceptibility to breast cancer due to CHEK2(*)1100delC in noncarriers of BRCA1 or BRCA2 mutations. Nat Genet, 2002. 31(1): p. 55–9.

12. Evans, D.G., et al., Penetrance estimates for BRCA1 and BRCA2 based on genetic testing in a Clinical Cancer Genetics service setting: risks of breast/ovarian cancer quoted should reflect the cancer burden in the family. BMC Cancer, 2008. 8: p. 155.

13. Minikel, E.V., et al., Ascertainment bias causes false signal of anticipation in genetic prion disease. Am J Hum Genet, 2014. 95(4): p. 371–82.

14. Turner, H. and L. Jackson, Evidence for penetrance in patients without a family history of disease: a systematic review. Eur J Hum Genet, 2020. 28(5): p. 539–550.

15. Easton, D.F., D. Ford, and D.T. Bishop, Breast and ovarian cancer incidence in BRCA1-mutation carriers. Breast Cancer Linkage Consortium. Am J Hum Genet, 1995. 56(1): p. 265–71.

16. Gabai-Kapara, E., et al., Population-based screening for breast and ovarian cancer risk due to BRCA1 and BRCA2. Proc Natl Acad Sci U S A, 2014. 111(39): p. 14205–10.

17. Abul-Husn, N.S., et al., Exome sequencing reveals a high prevalence of BRCA1 and BRCA2 founder variants in a diverse population-based biobank. Genome Medicine, 2019. 12(1): p. 2.

18. Grzymski, J.J., et al., Population genetic screening efficiently identifies carriers of autosomal dominant diseases. Nat Med, 2020. 26(8): p. 1235–1239.

19. Buchanan, A.H., et al., Clinical outcomes of a genomic screening program for actionable genetic conditions. Genet Med, 2020.

20. Centers for Disease Control and Prevention, Genomic Application Toolkit. 2019; Available from: https://www.cdc.gov/genomics/implementation/toolkit/tier1.htm.

21. National Comprehensive Cancer Network, Genetic/Familial High-Risk Assessment: Breast, Ovarian, and Pancreatic. 2020; Avarilable from: https://www.nccn.org/professionals/physician_gls/pdf/genetics_bop.pdf.

22. The eMERGE Consortium, Harmonizing Clinical Sequencing and Interpretation for the eMERGE III Network. Am J Hum Genet, 2019. 105(3): p. 588–605.

23. Richards, S., et al., Standards and guidelines for the interpretation of sequence variants: a joint consensus recommendation of the American College of Medical Genetics and Genomics and the Association for Molecular Pathology. Genet Med, 2015. 17(5): p. 405–24.

24. Riggs, E.R., et al., Technical standards for the interpretation and reporting of constitutional copy-number variants: a joint consensus recommendation of the American College of Medical Genetics and Genomics (ACMG) and the Clinical Genome Resource (ClinGen). Genet Med, 2020. 22(2): p. 245–257.

25. Wiesner, G.L., et al., Returning Results in the Genomic Era: Initial Experiences of the eMERGE Network. J Pers Med, 2020. 10(2).

26. Fossey, R., et al., Ethical Considerations Related to Return of Results from Genomic Medicine Projects: The eMERGE Network (Phase III) Experience. J Pers Med, 2018. 8(1).

27. Harris, P.A., et al., The REDCap consortium: Building an international community of software platform partners. J Biomed Inform, 2019. 95: p. 103208.

28. Harris, P.A., et al., Research electronic data capture (REDCap)--a metadata-driven methodology and workflow process for providing translational research informatics support. J Biomed Inform, 2009. 42(2): p. 377–81.

29. Kaplan, E.L. and P. Meier, Nonparametric Estimation from Incomplete Observations. Journal of the American Statistical Association, 1958. 53(282): p. 457–481.

30. Greenwood, M., A Report on the Natural Duration of Cancer, in Ministry of Health Reports on Public Health and Medical Subjects. 1926: London. p. 1–26.

31. Hao, J., et al., Healthcare Utilization and Costs after Receiving a Positive BRCA1/2 Result from a Genomic Screening Program. J Pers Med, 2020. 10(1).

32. Marabelli, M., S.C. Cheng, and G. Parmigiani, Penetrance of ATM Gene Mutations in Breast Cancer: A Meta-Analysis of Different Measures of Risk. Genet Epidemiol, 2016. 40(5): p. 425–31.

33. Maxwell, K.N., et al., Population Frequency of Germline BRCA1/2 Mutations. Journal of Clinical Oncology, 2016. 34(34): p. 4183–4185.

34. Laitman, Y., et al., Germline CHEK2 mutations in Jewish Ashkenazi women at high risk for breast cancer. Isr Med Assoc J, 2007. 9(11): p. 791–6.

35. Leedom, T.P., et al., Breast cancer risk is similar for CHEK2 founder and non-founder mutation carriers. Cancer Genet, 2016. 209(9): p. 403–407.

36. Gronwald, J., et al., Cancer risks in first-degree relatives of CHEK2 mutation carriers: effects of mutation type and cancer site in proband. Br J Cancer, 2009. 100(9): p. 1508–12.

37. Antoniou, A.C., et al., Breast-cancer risk in families with mutations in PALB2. N Engl J Med, 2014. 371(6): p. 497–506.

38. Amadou, A., M.I.W. Achatz, and P. Hainaut, Revisiting tumor patterns and penetrance in germline TP53 mutation carriers: temporal phases of Li-Fraumeni syndrome. Curr Opin Oncol, 2018. 30(1): p. 23–29.

39. Cancer Statistics Center, Probability of developing cancer, 2014-2016. Available from: https://cancerstatisticscenter.cancer.org/?_ga=2.226543582.276505911.1599136266-1567578018.1599136266#!/cancer-site/Breast.

40. National Comprehensive Cancer Network, Breast Cancer Risk Reduction. 2020; Version 1.2020:[Available from: https://www.nccn.org/professionals/physician_gls/pdf/breast_risk.pdf.

41. Network, N.C.C. Breast Cancer Screening and Diagnosis. 2019; Version 1.2019:[Available from: https://www.nccn.org/professionals/physician_gls/pdf/breast-screening.pdf.

42. Williams, M.S., et al., Patient-Centered Precision Health In A Learning Health Care System: Geisinger’s Genomic Medicine Experience. Health Aff (Millwood), 2018. 37(5): p. 757–764.

43. Han, M.R., et al., Evaluating genetic variants associated with breast cancer risk in high and moderate-penetrance genes in Asians. Carcinogenesis, 2017. 38(5): p. 511–518.

