## Supplement for "Penetrance of breast cancer genes from the eMERGE III Network"

**Supplementary Figures and Tables**

Xiao Fan, PhD^1, 2^, Julia Wynn, MS^1^, Ning Shang, PhD^3^, Cong Liu, PhD^3^, Alexander Fedotov, PhD^4^, Miranda L.G. Hallquist, MS^14^, Adam H. Buchanan, MS^14^, Marc S. Williams, MD^14^, Maureen E. Smith, MS^5^, Christin Hoell, MS^6^, Laura J. Rasmussen-Torvik, PhD^7^, Josh F. Peterson, MD^8^, Georgia L. Wiesner, MD^9^, Andrea M. Murad, MS^10^, Gail P. Jarvik, MD, PhD^11^, Adam S. Gordon, PhD^6^, Elisabeth A. Rosenthal, MD^11^, Ian B. Stanaway, PhD^11^, David R. Crosslin, PhD^12^, Eric B. Larson, MD^13^, Kathleen A. Leppig, MD^13^, Nora B. Henrikson, PhD^13^, Janet L. Williams, MS^14^, Rongling Li, PhD^15^, Scott Hebbring, PhD^16^, Chunhua Weng, PhD^3^, Yufeng Shen, PhD^2, 3,19^, Katherine D. Crew, MD^17,18,19^, Wendy K. Chung, MD, PhD^1, 17,18,19 *^

^1^ Department of Pediatrics, Columbia University Irving Medical Center, New York, NY 10032

^2^ Department of Systems Biology, Columbia University Irving Medical Center, New York, NY 10032

^3^ Department of Biomedical Informatics, Columbia University Irving Medical Center, New York, NY 10032

^4^ Irving Institute for Clinical and Translational Research, Columbia University Irving Medical Center, New York, NY 10032

^5^ Department of Medicine, Northwestern University, Chicago Feinberg School of Medicine, IL 60657

^6^ Center for Genetic Medicine, Northwestern University Feinberg School of Medicine, Chicago, IL 60611

^7^ Department of Preventive Medicine, Northwestern University Feinberg School of Medicine, Chicago, IL 60611

^8^ Department of Biomedical Informatics, Vanderbilt University Medical Center, Nashville TN 37203

^9^ Department of Medicine, Vanderbilt University Medical Center, Nashville, TN 37203

^10^ Division of Genetic Medicine, Department of Internal Medicine, University of Michigan, Ann Arbor, MI 48109

^11^ Department of Medicine (Medical Genetics), University of Washington Medical Center, Seattle, WA, 98105

^12^ Department of Biomedical Informatics and Medical Education, University of Washington Medical Center, Seattle, WA, 98105

^13^ Kaiser Permanente of Washington, Seattle, WA 98112

^14^ Genomic Medicine Institute, Geisinger, Danville, PA 17822

^15^ Division of Genomic Medicine, National Human Genome Research Institute, National Institutes of Health, Baltimore, MD 21224

^16^ Center for Precision Medicine Research, Marshfield Clinic, Marshfield, WI 54449

^17^ Herbert Irving Comprehensive Cancer Center, Columbia University Irving Medical Center, New York, NY 10032

^18^ Department of Medicine, Columbia University Irving Medical Center, New York, NY 10032

^19^ These authors jointly supervised this work

number of individuals

current age (years)

|  | not developed BC | developed BC | prophylactic surgeries |
| --- | --- | --- | --- |
| ATM | 15 | 6 | 0 |
| BRCA1 | 13 | 3 | 1 |
| BRCA2 | 32 | 6 | 1 |
| CHEK2 | 36 | 11 | 1 |
| PALB2 | 11 | 4 | 0 |
| PTEN | 3 | 0 | 0 |
| TP53 | 3 | 2 | 0 |

|  | current age |
| --- | --- |
| 1 | 63.1905 |
| 2 | 43.1765 |
| 3 | 50.3077 |
| 4 | 59.4167 |
| 5 | 61.4 |
| 6 | 32 |
| 7 | 51.2 |

**Supplementary Figure S1. Breast cancer (BC) status by the last review including not developed breast cancer (blue), developed breast cancer (orange) and prophylactic mastectomy (red). The average age and standard deviation were shown in the secondary y axis.**

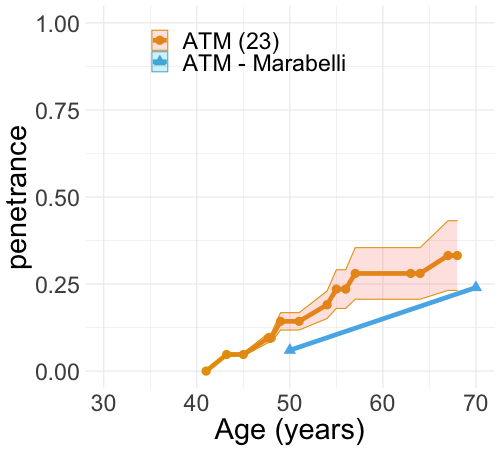

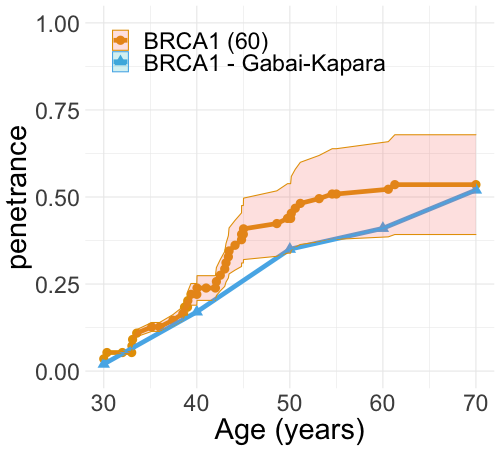

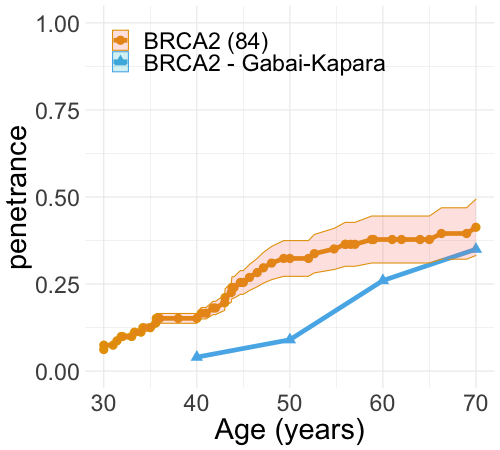

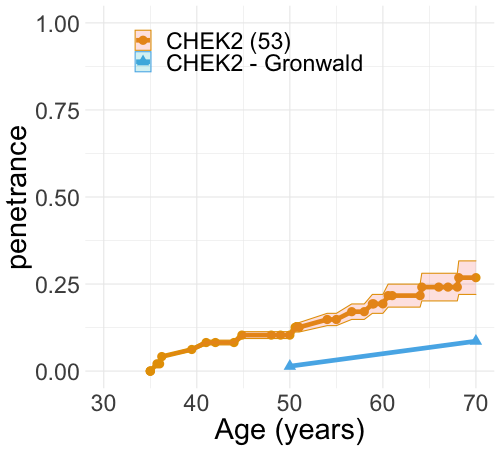

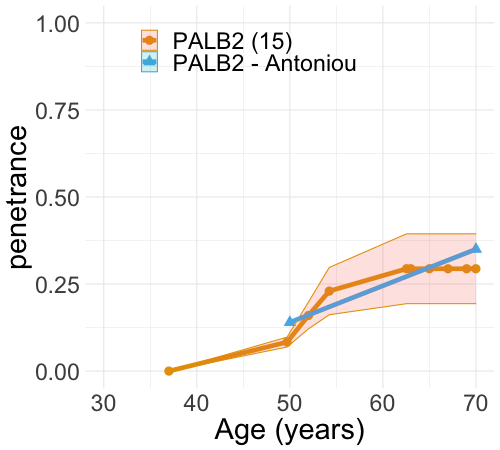

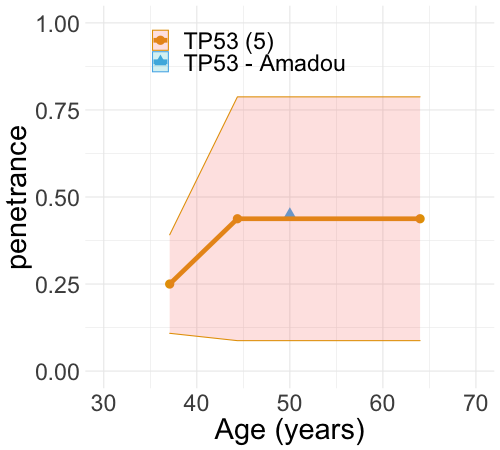

**Supplementary Figure S2. Penetrance (orange) of six breast cancer genes compared with penetrance (blue) in the literature from Table 2. The sample size of the entire eMERGE III including those who were aware of their breast cancer genetic results prior to enrollment in eMERGE. Orange bands indicate 95% confidence intervals. Genes are sorted alphabetically.**

**Supplementary Table S1. Clinical characteristics of 24,947 eMERGE III participants.**

The sex was genetically inferred and the ancestry was self-reported. The age is current age.

| sex | ancestry | count | age (years) |
| --- | --- | --- | --- |
| female (N=13,458) | AFR | 1758 | 38.6±23.5 |
|  | AMR | 960 | 49.5±18 |
|  | EAS | 952 | 52.3±14.2 |
|  | EUR | 9122 | 54.1±21.3 |
|  | SAS | 65 | 63.1±13.9 |
|  | unknown | 601 | 48.8±23.1 |
| male (N=11,498) | AFR | 1994 | 31.5±21.8 |
|  | AMR | 546 | 46±20.6 |
|  | EAS | 588 | 53.9±16 |
|  | EUR | 7892 | 54.1±23.8 |
|  | SAS | 34 | 51.3±20.1 |
|  | unknown | 444 | 41.3±24.7 |

**Supplementary Table S2. Recurrent P/LP variants in the breast cancer susceptibility genes, sorted by allele count in the eMERGE penetrance cohort.**

| cDNA | Gene | Function | ClinVar | Allele count |
| --- | --- | --- | --- | --- |
| c.470T>C | *CHEK2* | missense | Conflicting interpretations of pathogenicity | 26 |
| c.1100delC | *CHEK2* | frameshift | Conflicting interpretations of pathogenicity | 7 |
| c.2638+2T>C | *ATM* | splice-donor | Pathogenic/Likely pathogenic by multiple submitters | 4 |
| c.5946del | *BRCA2* | frameshift | Pathogenic reviewed by expert panel | 3 |
| c.1427C>T | *CHEK2* | missense | Conflicting interpretations of pathogenicity | 3 |
| c.1283C>T | *CHEK2* | missense | Conflicting interpretations of pathogenicity | 3 |
| c.7271T>G | *ATM* | missense | Pathogenic/Likely pathogenic by multiple submitters | 2 |
| c.3049C>T | *ATM* | stop-gain | Pathogenic by multiple submitters | 2 |
| c.4876_4877delAA | *BRCA2* | frameshift | Pathogenic reviewed by expert panel | 2 |
| c.5217_5223delTTTAAGT | *BRCA2* | frameshift | Pathogenic reviewed by expert panel | 2 |
| c.9253dupA | *BRCA2* | frameshift | Pathogenic reviewed by expert panel | 2 |
| c.2808_2811delACAA | *BRCA2* | frameshift | Pathogenic reviewed by expert panel | 2 |
| c.7558C>T | *BRCA2* | stop-gain | Pathogenic reviewed by expert panel | 2 |
| c.2257C>T | *PALB2* | stop-gain | Pathogenic by multiple submitters | 2 |

**Supplementary Table S3. Number of carriers and unique P/LP variants for each gene and variant type. Number of variants with at least 2-star review status in ClinVar is given in the last column with percentage in the parentheses.**

|  | Number of carriers | | | | | |
| --- | --- | --- | --- | --- | --- | --- |
| gene | missense | splice | stop-gain | frameshift | CNV | ClinVar |
| *ATM* | 3 | 6 | 7 | 5 | 0 | 18 (86%) |
| *BRCA1* | 3 | 2 | 4 | 8 | 0 | 15 (88%) |
| *BRCA2* | 0 | 5 | 8 | 26 | 0 | 37 (95%) |
| *CHEK2* | 33 | 2 | 1 | 10 | 2 | 7 (15%) |
| *PALB2* | 0 | 4 | 6 | 5 | 0 | 14 (93%) |
| *PTEN* | 1 | 0 | 1 | 1 | 0 | 3 (100%) |
| *TP53* | 3 | 0 | 2 | 0 | 0 | 4 (80%) |
|  | Numbers of unique variants | | | | | |
| gene | missense | splice | stop-gain | frameshift | CNV | ClinVar |
| *ATM* | 2 | 3 | 6 | 5 | 0 | 13 (81%) |
| *BRCA1* | 3 | 2 | 4 | 8 | 0 | 15 (88%) |
| *BRCA2* | 0 | 5 | 7 | 20 | 0 | 30 (94%) |
| *CHEK2* | 4 | 2 | 1 | 4 | 2 | 7 (54%) |
| *PALB2* | 0 | 4 | 5 | 5 | 0 | 13 (93%) |
| *PTEN* | 1 | 0 | 1 | 1 | 0 | 3 (100%) |
| *TP53* | 3 | 0 | 2 | 0 | 0 | 4 (80%) |

CNV is copy number variation.

**Supplementary Table S4. Summary of recent penetrance studies that were used to compare with these population-based penetrance estimates for the six breast cancer genes.**

| Genes | Number of women with P/LP variants | Source of the study populations | Ancestry | Reference |
| --- | --- | --- | --- | --- |
| *ATM* | NA | Meta case-control studies | multiple | [1] |
| *BRCA1/2* | 211 | General population | Ashkenazi Jewish | [2] |
| *CHEK2* | 533 | Cancer clinic | European | [3] |
| *PALB2* | 311 | Cancer clinic | European | [4] |
| *PTEN* | 368 | Breast Cancer Association Consortium | Asian | [5] |
| *TP53* | 2079 | International Agency for Research on Cancer | multiple | [6] |

**References:**

1.	Marabelli, M., S.C. Cheng, and G. Parmigiani, *Penetrance of ATM Gene Mutations in Breast Cancer: A Meta-Analysis of Different Measures of Risk.* Genet Epidemiol, 2016. **40**(5): p. 425-31.

2.	Gabai-Kapara, E., et al., *Population-based screening for breast and ovarian cancer risk due to BRCA1 and BRCA2.* Proc Natl Acad Sci U S A, 2014. **111**(39): p. 14205-10.

3.	Gronwald, J., et al., *Cancer risks in first-degree relatives of CHEK2 mutation carriers: effects of mutation type and cancer site in proband.* Br J Cancer, 2009. **100**(9): p. 1508-12.

4.	Antoniou, A.C., et al., *Breast-cancer risk in families with mutations in PALB2.* N Engl J Med, 2014. **371**(6): p. 497-506.

5.	Han, M.R., et al., *Evaluating genetic variants associated with breast cancer risk in high and moderate-penetrance genes in Asians.* Carcinogenesis, 2017. **38**(5): p. 511-518.

6.	Amadou, A., M.I.W. Achatz, and P. Hainaut, *Revisiting tumor patterns and penetrance in germline TP53 mutation carriers: temporal phases of Li-Fraumeni syndrome.* Curr Opin Oncol, 2018. **30**(1): p. 23-29.
